# Re-use of trial data in the first 10 years of the data-sharing policy of the Annals of Internal Medicine: a survey of published studies

**DOI:** 10.1101/2020.06.16.20132894

**Authors:** Claude Pellen, Laura Caquelin, Alexia Jouvance-Le Bail, Jeanne Gaba, Mathilde Vérin, David Moher, John P. A. Ioannidis, Florian Naudet

## Abstract

**Background:** The Annals of Internal Medicine (AIM) has adopted a policy encouraging data-sharing since 2007.

**Objective:** To explore the impact of the AIM data-sharing policy for randomized controlled trials (RCTs) in terms of output from data-sharing (i.e. publications re-using the data).

**Design:** Retrospective study.

**Setting:** AIM.

**Participants:** RCTs published in the AIM between 2007 and 2017 were retrieved on PubMed. Publications where the data had been re-used were identified on Web of Science. Searches were performed by two independent reviewers.

**Interventions:** Intention to share data (or not) expressed in a data-sharing statement.

**Measurements:** The primary outcome was any published re-use of the data (i.e. re-analysis, secondary analysis, or meta-analysis of individual participant data [MIPD]), where the first, last and corresponding authors were not among the authors of the RCT. Components of the primary outcome and analyses without any author restriction were secondary outcomes. Analyses used Cox (primary analysis) models adjusting for RCT characteristics.

**Results:** 185 RCTs were identified. 106 (57%) mentioned willingness to share data and 79 (43%) did not. 208 secondary analyses, 67 MIPD and no re-analyses were identified. No significant association was found between intent to share and re-use where the first, last and corresponding authors were not among the authors of the primary RCT (adjusted hazard ratio = 1.04 [0.47-2.30]). Secondary outcomes also showed no association between intent to share and re-use.

**Limitations:** Possibility of residual confounding and limited power.

**Conclusion:** Over ten years, RCTs published in AIM expressing an intention to share data were not associated with more extensive re-use of the data.

**Registration:** https://osf.io/8pj5e/

**Funding Source:** Grants from the Fondation pour la Recherche Médicale, Région Bretagne, and French National Research Agency.

## INTRODUCTION

Data-sharing (i.e. sharing of data, codes, programs and material) is the norm in many scientific disciplines, but until recently this has not been the case with biomedical research (2). In medicine, randomized controlled trials (RCTs) are considered influential and therefore availability of their data is expected to be crucial in the evaluation of health interventions (e.g. for meta-analyses of individual participant data, MIPD). In June 2017, the International Committee of Medical Journal Editors (ICMJE) required a data-sharing plan to be included in each paper (and pre-specified in study registration) (3). As these new requirements for publishing experimental findings come into effect, it is necessary to assess whether they have their intended effects.

Among the leading general medical journals, the New England Journal of Medicine (NEJM), The Lancet, the JAMA and JAMA Internal Medicine have had no specific policy for sharing data from RCTs until recently. The British Medical Journal (BMJ) and the Public Library of Science (PLOS) Medicine have already adopted stronger policies, reaching beyond the ICMJE policy, which render data-sharing mandatory with the publication of RCTs. Nevertheless, in view of actual data-sharing rates their policy seems imperfect (4).

The Annals of Internal Medicine (AIM) has encouraged (but not required) data-sharing since 2007 (5). Since then, the journal has required a reproducible research statement to be included in every original research article (including RCTs). This reproducible research statement indicates “whether the study protocol, data, or statistical code is available to readers, and under what terms authors will share this information.” While this policy did not make data-sharing mandatory, its aim was “to help the scientific community evaluate, and build upon, the research findings” published in AIM. Importantly, this is to a large extent what is required by the new ICMJE policy. Therefore, a retrospective analysis of RCTs published in AIM between 2007 and 2017 could provide a proxy for the expected impact of the ICMJE policy.

We explored the effectiveness of RCT data-sharing from AIM publications in terms of output from data-sharing (i.e. publications where the data has been re-used). We specifically aimed to describe the data-sharing practices in RCTs published in AIM over a decade (2007-2017), to assess the association between intent to share and published re-uses of data, and to assess the association of intent to share with citation rates.

## METHODS

The methods were specified in advance. They were documented in a protocol registered with the Open Science Framework (OSF) on 3rd August 2018 (https://osf.io/gnt6u/).

### Eligibility criteria

We surveyed a retrospective cohort of RCTs published in the AIM between April 2007 and December 2017. The RCTs included the following designs: two parallel groups and multiple groups, cluster trials and cross-over studies, non-inferiority and superiority trials. All publications were inspected to exclude secondary analyses and re-analyses of a previously published RCT. Publications without reproducible research statements (i.e. those that did not comply with the policy) were excluded.

### Search strategy and selection of primary publications of studies in AIM

We identified eligible studies from PubMed/Medline using the following strategy: (annals of internal medicine) AND (“2007/04/01”[Date - Publication]: “2017/12/31”[Date - Publication]) / limitation: randomized controlled trial.

Two reviewers (CP and AJLB) performed the eligibility assessment independently. Disagreements were resolved by consensus or in consultation with a third reviewer (FN).

### Search strategy and study selection for published re-uses

We used the Clarivate Web of Science database to identify secondary publications derived from these primary trials, since it would be extremely unlikely for a secondary publication not to cite the primary trial. We identified and recorded the total number of citations for each primary publication. We recorded the number of citations by articles that used individual-level data from the primary article in the Annals. For articles considered as potentially secondary publications, the abstracts and, when necessary, full texts were examined by two independent reviewers (among LC, JG, and MV) to confirm eligibility. One reviewer inspected all included citations and when he disagreed with the inclusion, disagreements were resolved in consultation with a third reviewer (FN). In addition, whenever a methodological article was cited in the primary article in the Annals (i.e. an article describing the methods and protocol of a trial), we entered this article in the Web of Science searches to identify additional citations.

### Data extraction

A data extraction sheet was developed. For each article included, we extracted information on study characteristics (date of publication, country (USA/Europe/Asia/other), intervention type (drug/device/complex intervention), control group (active/inactive), medical specialty (medicine/surgery/psychiatry), total sample size, result on the primary outcome (positive/negative), and funding source (academic/industry/charity/mixed). Detailed information on the data-sharing plan was extracted from the reproducible research statement. We recorded whether a statement indicated that the data was available (i.e. intent to share data). If the authors intended to share data, we extracted the type of data-sharing plan (1/ directly available, 2/ available upon request) and the type of material that was intended to be shared (1/ data-set, 2/ code, 3/ study protocol). If the authors did not intend to share data, we extracted reasons for not sharing data (when available).

Two authors (CP and AJLB) independently extracted the data from the studies included. Disagreements were resolved by consensus or in consultation with a third reviewer (FN).

### Outcomes

Our primary outcome was the re-use of data (yes/no) documented in all the citing articles on Web of Science. This is a composite outcome defined by any secondary use 1/ in a re-analysis, 2/ in a secondary analysis and 3/ in a MIPD (including pooled analyses without systematic review). The primary outcome was limited to published re-uses where the first, last and corresponding authors were not among the authors of the primary article in the Annals.

The pre-specified secondary outcomes were as follows: 1/ Components (1, 2, and 3) of the primary outcome; 2/ Re-use of the data (yes/no) documented in all the citing articles, on Web of Science without the author restriction used for the primary outcome; 3/ Number and type of secondary use (re-analyses, secondary analyses, MIPD) in which the lead authors (first, corresponding and last) of the published re-use of the data were outside the team authoring the primary article in the Annals; 4/ Number and type of secondary use (re-analyses, secondary analyses, MIPD) by an independent team (no author in common); 5/ Number and type of secondary use (re-analyses, secondary analyses, MIPD) by a team where <50% of authors were among the authors of the primary article in the Annals; 6/ Number and type of secondary use (re-analyses, secondary analyses, MIPD) by a team where ≥50% of authors were among the authors of the primary article in the Annals; 7/ Number and type of secondary use (re-analyses, secondary analyses, MIPD) by a team where 100% of authors were among the authors of the primary article in the Annals; 8/ Mention in the published re-uses of the availability of the data re-used (yes/no) and how it was used (qualitative data); 9/ Number of citations (on Web of Science).

### Statistical analysis

A detailed statistical analysis plan was registered with the OSF on 1^st^ August 2019 (https://osf.io/zck9y/) prior to merging the AIM primary articles database and the re-use database. The general strategy for modeling was as follows: we first used a univariate model with fixed effects to explore solely the relationship between the dependent and the explanatory variables, adjusting on possible confounders (country, intervention type, control group, medical specialty, result (positive/negative) on the primary outcome, funding source, and sample size). All associations with a p-value <0.25 were subsequently explored in a multivariate model. In all final models, we considered adding a random effect to account for studies that were published by the same team (defined as groups of primary AIM articles clustered by any common authors).

We described ‘intent to share data’ rates over time using a logistic regression. Results are expressed as odds ratios (OR) with their 95% confidence intervals (CIs). The primary outcome and all its components were analyzed using Kaplan-Meier and compared using proportional hazards regression. Results are expressed as hazard ratios (HR) with their 95% CI. Other secondary outcomes were analyzed as count data using Poisson or (when necessary) binomial negative regressions. Results are expressed as incidence rate ratios (IRR) with their 95% CI.

All statistical analyses were performed at a 5% significance level (two-sided tests). All analyses were performed under R version 3.4.1 (2017-06-30, The R Foundation for Statistical Computing).

### Changes from the initial protocol

The following clarifications were described prior to the analyses in the statistical analysis plan: 1/ we decided to classify pooled analyses as MIPD because this point was unclear in the initial protocol, 2/ we made it clear that the year of publication (and date) was used to calculate each follow-up time (date of publication to date of citation searches) in survival models and as an offset in count models, and not as a potential confounder and 3/ also, we excluded citations concerning long-term follow-up and use of stored biological samples, since these analyses obviously implied the acquisition of new data.

We considered the sample size of the primary article in the Annals as a potential confounder, since it could be responsible for changes in both the intent to share and the number of re-uses, and therefore could be an important confounding factor, which we omitted to plan for in our initial protocol.

While we initially planned to include 3 types of re-use (secondary analyses, re-analyses and MIPD), we decided to include meta-analyses of aggregate data as supplementary analyses. Indeed, data availability can have an impact when the initial publication does not report the data required to perform the meta-analysis.

## RESULTS

### Characteristics of primary articles and published re-uses

**Figure 1** shows the study selection process. The searches carried out on 27^th^ May 2018 retrieved 257 items. Of these, 185 were primary articles and had a data-sharing statement; these articles were included. These RCTs had a median sample size of 299 participants (interquartile range (IQR) [158-719]), they had academic funding in 79 cases (42.7%), and were led by teams from USA in 97 cases (52.4%). Eighty-eight RCTs (47.6%) evaluated drug interventions, 89 (48.1%) complex interventions (e.g. psychotherapeutic program), and 8 (4.3 %) evaluated devices. 106 studies mentioned willingness to share the dataset and 79 mentioned no intent to share their dataset. Of these, none provided a reason for not sharing. **Table 1** presents the characteristics of these studies displayed by intent to share or not.

**Table 1.**
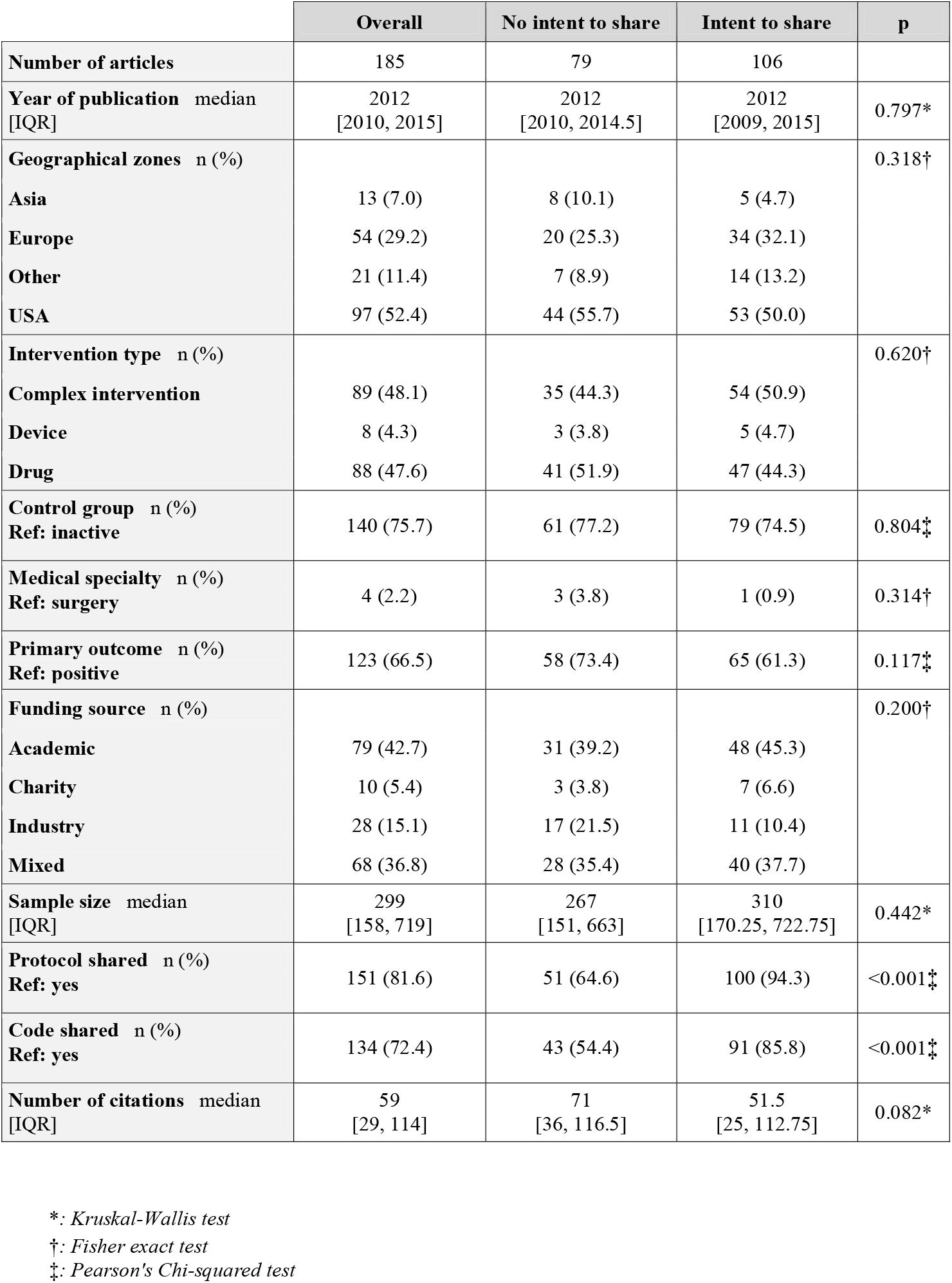
Characteristics of the randomized controlled trials included.

|  | Overall | No intent to share | Intent to share | p |
| --- | --- | --- | --- | --- |
| **Number of articles** | 185 | 79 | 106 |  |
| **Year of publication** median [IQR] | 2012 [2010, 2015] | 2012 [2010, 2014.5] | 2012 [2009, 2015] | 0.797* |
| **Geographical zones** n (%) |  |  |  | 0.318† |
| Asia | 13 (7.0) | 8 (10.1) | 5 (4.7) |  |
| Europe | 54 (29.2) | 20 (25.3) | 34 (32.1) |  |
| Other | 21 (11.4) | 7 (8.9) | 14 (13.2) |  |
| USA | 97 (52.4) | 44 (55.7) | 53 (50.0) |  |
| **Intervention type** n (%) |  |  |  | 0.620† |
| Complex intervention | 89 (48.1) | 35 (44.3) | 54 (50.9) |  |
| Device | 8 (4.3) | 3 (3.8) | 5 (4.7) |  |
| Drug | 88 (47.6) | 41 (51.9) | 47 (44.3) |  |
| **Control group** n (%)<br>**Ref: inactive** | 140 (75.7) | 61 (77.2) | 79 (74.5) | 0.804‡ |
| **Medical specialty** n (%)<br>**Ref: surgery** | 4 (2.2) | 3 (3.8) | 1 (0.9) | 0.314† |
| **Primary outcome** n (%)<br>**Ref: positive** | 123 (66.5) | 58 (73.4) | 65 (61.3) | 0.117‡ |
| **Funding source** n (%) |  |  |  | 0.200† |
| Academic | 79 (42.7) | 31 (39.2) | 48 (45.3) |  |
| Charity | 10 (5.4) | 3 (3.8) | 7 (6.6) |  |
| Industry | 28 (15.1) | 17 (21.5) | 11 (10.4) |  |
| Mixed | 68 (36.8) | 28 (35.4) | 40 (37.7) |  |
| **Sample size** median [IQR] | 299 [158, 719] | 267 [151, 663] | 310 [170.25, 722.75] | 0.442* |
| **Protocol shared** n (%)<br>**Ref: yes** | 151 (81.6) | 51 (64.6) | 100 (94.3) | <0.001‡ |
| **Code shared** n (%)<br>**Ref: yes** | 134 (72.4) | 43 (54.4) | 91 (85.8) | <0.001‡ |
| **Number of citations** median [IQR] | 59 [29, 114] | 71 [36, 116.5] | 51.5 [25, 112.75] | 0.082* |

\*: Kruskal-Wallis test
†: Fisher exact test
‡: Pearson's Chi-squared test

**Figure 1.**
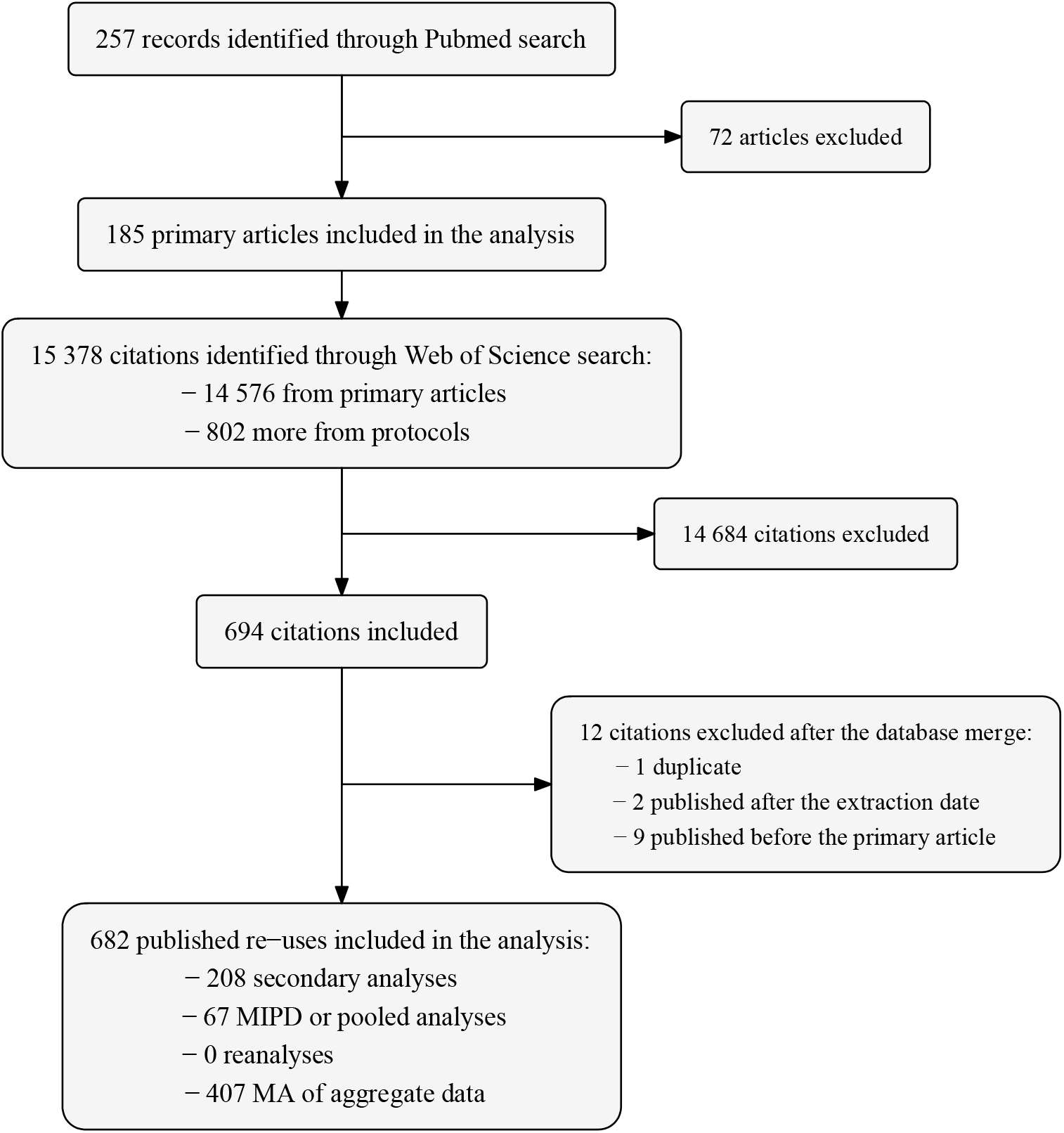
Study flow diagram.

Search for re-uses of these primary articles were carried out on 9^th^ August 2018. There were 15,378 citations identified through Web of Science searches (14,576 from primary articles and 802 more from protocols of these studies). Among these, we identified 208 secondary analyses, 67 MIPD, and 0 re-analyses. We also identified 407 meta-analyses of aggregate data.

### Intent to share trends over time

**Figure 2** shows data-sharing trends over time. We found no association between ‘intent to share data’ rates and time (years) (OR = 1.00 [0.90 −1.11], adjusted OR = 0.98 [0.84-1.13]). Details of the adjusted model are provided in **Web-appendix 1**. Proportions of code and protocol availability were always higher than levels of intent to share data, and reached 100 % for protocol availability in 2017.

**Figure 2.**
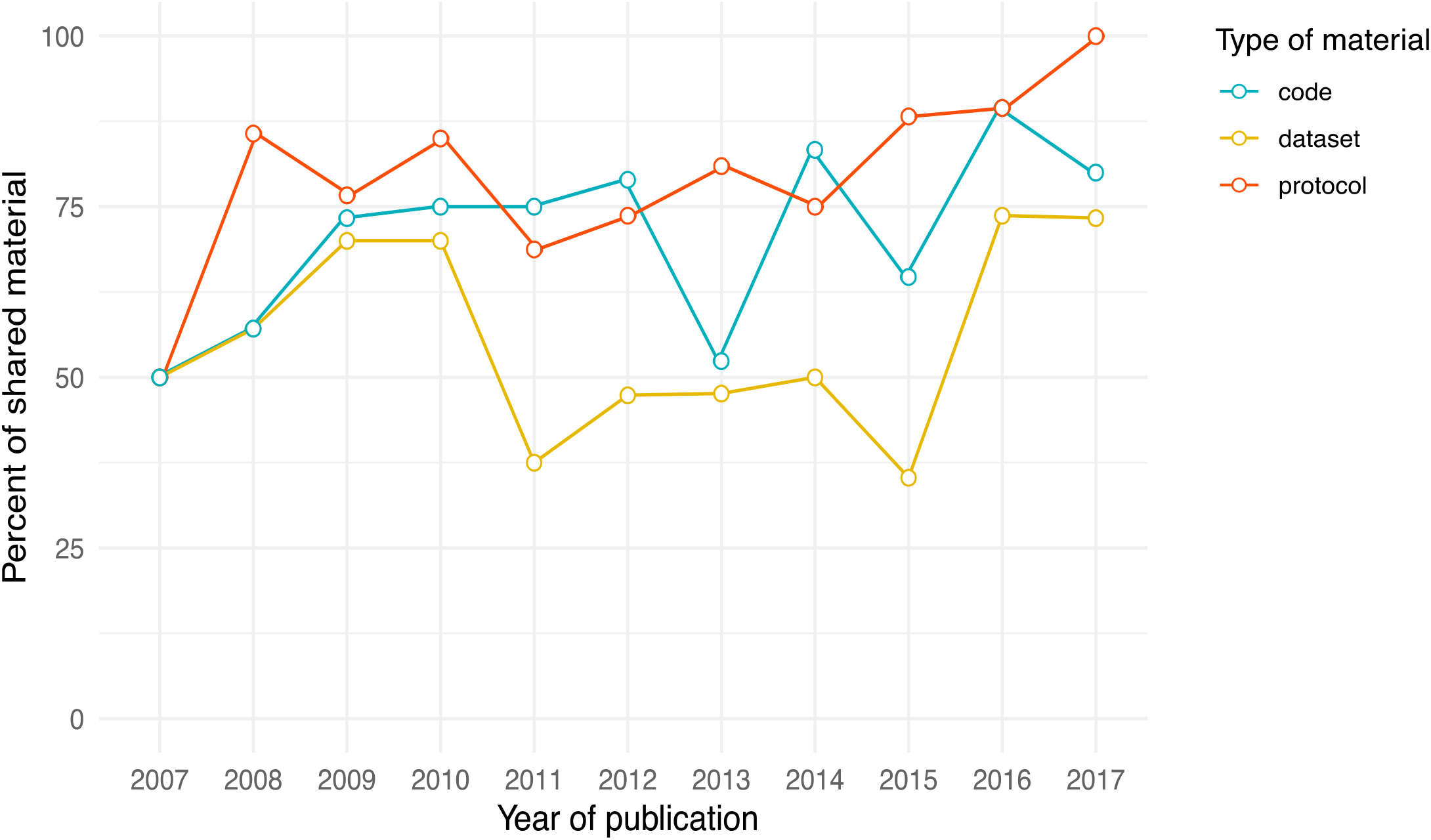
Data-sharing rates for protocol, statistical code and dataset of RCTs published in the Annals of Internal Medicine over time.

### Association between intent to share and published re-uses of data

Univariate and multivariate analyses (**Figure 3**) identified no significant association between intent to share and the primary outcome, that is to say published re-uses of the data where the first, last and corresponding authors were not among the authors of the primary article in the Annals (adjusted HR = 1.04 [0.47-2.30]), nor was there any association with its different components (adjusted HR = 0.96 [0.32-2.90] for secondary analyses, adjusted HR = 1.23 [0.37-4.06] for MIPD). The same results were observed in analyses using no author restriction criteria (adjusted HR = 1.30 [0.84-2.01]). Supplementary analyses, including meta-analyses of aggregate data not specified in the registered protocol, are presented in **Web-appendix 2**. The results were consistent with those of the main analysis.

**Figure 3.**
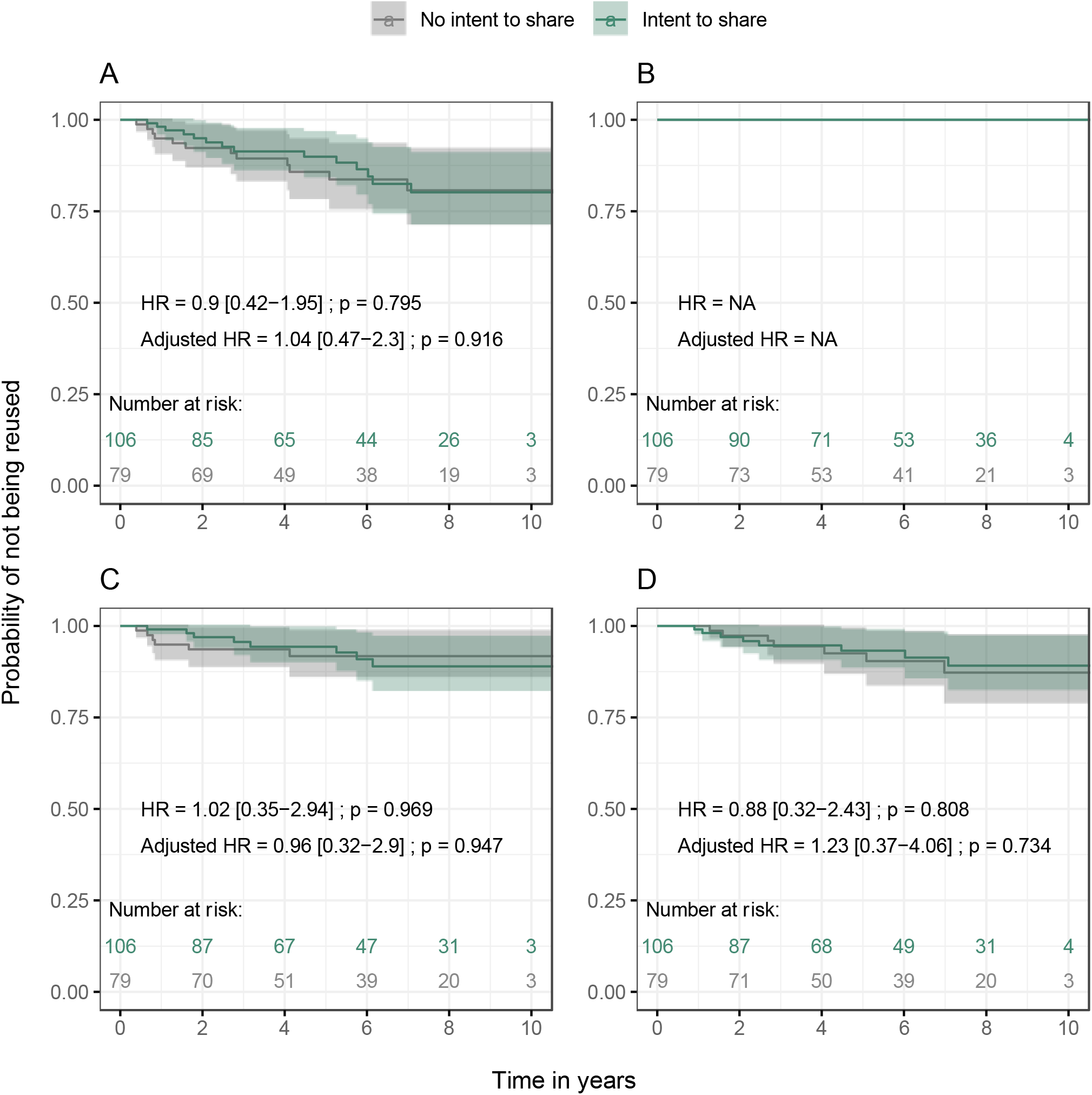
Kaplan-Meier curves and hazard ratios (HR) of published re-uses where the first, last and corresponding authors are not among the authors of the primary article. *A: any type of published re-uses. B: published re-analyses. C: published secondary analyses. D: published MIPD*.

**Figure 4** presents count outcomes related to the number of re-uses (by different types of reuse). A few statistically significant associations were found in univariate analyses but they were not confirmed in multivariate analyses for pre-specified outcomes. Only the outcome “MIPD/pooled analyses with no author in common with the primary RCT” retained a weak association signal in multivariate analyses. The number tended to be greater when there was no mention of data-sharing (adjusted IRR = 0.03 [0.00-0.33]). A careful examination of individual papers found that among the 13 re-uses of RCTs stating they have no intention to share, 12 had the same funder as the primary RCTs, all of these being sponsored by pharmaceutical firms (**Web-appendix 3**). These results are presented in detail in **Web-appendix 4**, and the number of re-uses per RCT are presented in **Web-appendix 5**.

**Figure 4.**
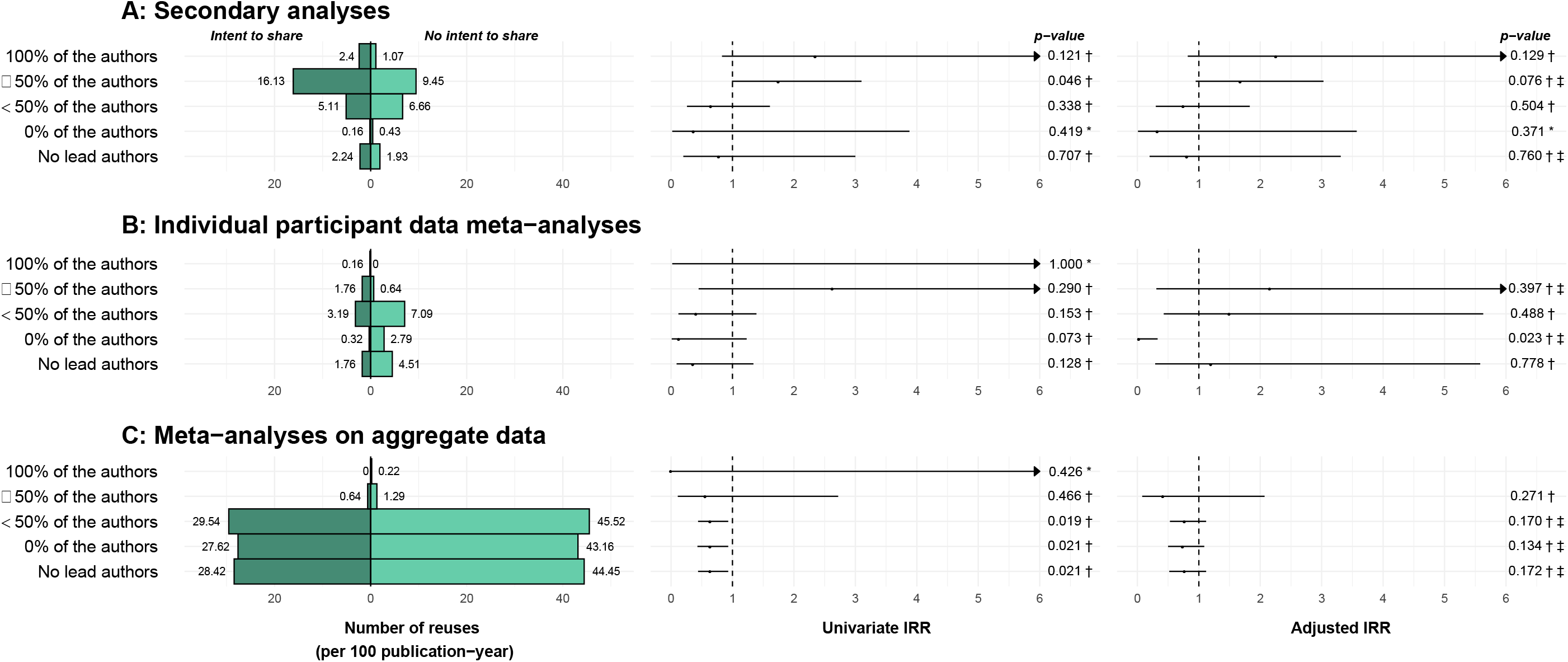
Number of re-uses by intent to share. *: *Poisson model* †: *Binomial negative model* ‡: *Mixed model*

### Association between intent to share and number of citations

Univariate and multivariate analyses identified no significant association between intent to share and the number of citations, with 134 citations per 10 publication-years for papers with an intention to share statement and 149 per 10 publication-years for papers stating they had no intention to share (IRR = 0.83 [0.67-1.02]; adjusted IRR = 0.90 [0.72-1.13]).

## DISCUSSION

### Statement of primary findings

Over ten years, one might have expected a progressive increase in data-sharing practices with a progressive improvement in data-sharing intent over time. However, and in line with a previous exploration of intent to share data for all research articles in AIM between 2008 and 2012 (6), we did not find any such increase. Interestingly, while Laine et al. (8) found that the intention to share protocols was lower than for the intention to share datasets, we found the opposite, with higher, increasing rates of intention to share protocols, reaching 100% in 2017. This result could be explained by our specific focus on clinical trials. Protocol availability is a core feature of transparency for these studies, especially in the context of the strong, justified external pressure by some watchdog groups. For instance, in response to the efforts by the Centre for Evidence-Based Medicine Outcome Monitoring Project (COMPare) (9) to document outcome switching in RCTs, the Annals editor recalled that the journal routinely asks authors of clinical trials to submit their protocols with their manuscripts, and examines trial registries for the initial and final information entered about trials (10). In data-sharing statements, intention to share codes was always more prominent than the sharing of data. This result is curious, since codes without the data are most often useless.

While more than 55% of the primary articles surveyed intended to make their data available, no association was found between intent to share and re-use of the data. Interestingly, there were only a few published re-uses of the primary articles, and three-quarters were secondary analyses. MIPD were the remaining re-uses and we found no re-analyses, meaning that none of the re-uses of the data was for reproducibility purposes. This last result is unfortunate given the increased deployment of reproducibility checks in other fields (11). However, this observation is in line with an earlier survey (12) exploring requests for data access from the National Heart, Lung and Blood Institute data repository. Over 16 years, 100 trials were available, 88 of them had a data request and 47 had at least one publication resulting from data-sharing. More than 80% of requests were for secondary analyses or methodological developments, and only 7% were for MIPD. In this survey, only two requests concerned re-analyses. And indeed, there is almost no culture of performing and publishing re-analyses in the clinical trial literature (13), especially by entirely independent authors. Another survey of cardiometabolic clinical trials available on the clinical study data request platform (14) suggested that despite efforts to make data available, re-uses were rare. Over 4 years, only 3 re-uses had been published among the 537 studies available for access. Similarly, over 11 years (15), only 14 re-uses (5 secondary analyses and 9 MIPD) were published using 51 clinical trial datasets available on the Data Share platform of the National Institute on Drug Abuse. Altogether, these results suggest that, despite some important efforts, data-sharing does not systematically achieve large numbers of published outputs.

Different factors could explain this finding. First, the available datasets may not be requested (and used) by external researchers. Second, primary authors may refuse to share their data as promised in their data-sharing statement. For instance, in our survey, only one publication had its dataset directly available on a repository, while all the others were available upon request. Similar rates were observed in a survey (8) of the Annals’ first year of the reproducible research statement policy, and in a second survey (6) accounting for articles from 2008 to 2012. When data is available upon request, data retrieval could be suboptimal as suggested by a previous survey of studies published in PLOS Medicine or BMJ under a strict data-sharing policy (4). However, among 90 authors of trials published between January 1, 2012 and March 1, 2016 in PLOS Medicine, The BMJ and the Annals (16), half of the respondents had a data-sharing plan (n = 49) and about one third reported they had received at least one data-sharing request, and very few of these were reported as being refused. It is therefore possible that data is in fact shared, but that re-use of this data will not translate into published output. A cross-sectional web-based survey (17) of the NIH central database repository found that only 67% of the reuses were published. In addition, shared datasets could serve for pedagogical purpose or for designing trials (e.g. sample size calculation) or any other activity that may not necessarily lead to any published output. However, it is also possible that data-sharing enables numerous secondary analyses to be run, among which only a few reach published posterity. This last hypothesis is of concern, because sub-optimal reporting and selective publication of these re-uses could lead to non-reproducible research (18).

### Limitations

Our results should be interpreted cautiously. The observational design of the study does not enable us to draw causal inferences between intent to share and research outputs from actual re-use of data. Confounding is a major issue in observational research, and there is no perfect way to handle it. Despite the adjustment of the analyses to various available factors that might influence the patterns of re-use, some factors could not be fully accounted for. For instance, we were not able to adjust for subtle variations that can arise across different medical subspecialties or topics. Also, we did not identify the corresponding author’s gender or career stage, while a junior investigator could be more aware of open science practices such as data-sharing. Therefore, this study is prone to residual confounding. In our study, the identification of published re-uses proved to be a difficult task, with the risk of missing some studies. We minimized this risk by using a dual, independent extraction.

Also, it was difficult to define independent re-uses objectively. For instance, the primary article authors could require to be among authors of the re-use as part of their data-sharing statement, even if they were not actively involved in the re-use. This could have introduced some classification bias in our study. For instance, the only signal we identified was for IPD meta-analyses (or pooled analyses) with no author in common with the primary RCT, which were more frequently RCTs without intention to share data. The topics of these RCTs were long-term efficacy of dapagliflozin for patients with diabetes mellitus, adalimumab in moderate to severe Hidradenitis suppurativa, tofacitinib in associations for patients with active rheumatoid arthritis, varenicline in smoking cessation, and the Grazoprevir-Elbasvir combination in Chronic Hepatitis C Virus. The RCT that shared their data was an equivalence trial among treatment-naive volunteers infected with HIV-1. All but one of these reuses were sponsored by the same funder as the original trial. It is therefore likely that a given funder decided not to release the data and to perform their own research agenda, including series of pooled analyses. Also, classification bias could be differential between studies stating they do or do not intent to share data. For this reason, we used a restriction on the lead authors (first, last and corresponding) for our primary outcome, and performed a series of sensitivity analyses with various definitions. Importantly, all these analyses yielded very similar results.

Most importantly, this retrospective study explores an early experience of data-sharing. The primary articles included in our study cover 10 years during which the AIM pioneered clinical data-sharing, with a few other journals. As a result, re-uses like MIPD needing more than only one study dataset could have been hard to perform, because the overall environment was unfavorable. Up to 2015 most MIPDs were incomplete, half retrieved less than 80% of the eligible IPD and retrieval rates across published MIPDs did not improve through these years (19). Improved access to data of a single study may be of limited interest in this environment. Therefore, extrapolation to the new ICMJE policy should be very cautious, as we are operating in a fast-changing landscape.

## CONCLUSION

Data-sharing policies are coming progressively into effect, and are intended to reform the way clinical research is performed. Our analysis of one of the earliest experiences in the Annals suggests that things are not that simple. While it is hard to extrapolate these findings directly to the new ICMJE policy, our results highlight the need to assess whether this new policy will achieve its intended effects. In our opinion, the ICMJE policy should have an evaluation component, which is currently lacking.

## Data Availability

Study protocol, statistical code, data set and web-appendixes: available on the OSF platform

https://osf.io/8pj5e/

## Acknowledgment

The authors would like to thank Bruno Falissard and Etienne Dantan.

## Reproducible Research Statement

Study protocol, statistical code, data set and web-appendixes: directly available on the OSF platform (https://osf.io/8pj5e/).

